# SARS-CoV-2 Infection Hospitalization Rate and Infection Fatality Rate among the Non-Congregant Population in Connecticut

**DOI:** 10.1101/2020.10.30.20223461

**Authors:** Shiwani Mahajan, César Caraballo, Shu-Xia Li, Claire Dong, Lian Chen, Sara K. Huston, Rajesh Srinivasan, Carrie A. Redlich, Albert I. Ko, Jeremy S. Faust, Howard P. Forman, Harlan M. Krumholz

## Abstract

**Importance:** COVID-19 case fatality and hospitalization rates, calculated using the number of confirmed cases of COVID-19, have been described widely in the literature. However, the number of infections confirmed by testing underestimates the total infections as it is biased based on the availability of testing and because asymptomatic individuals may remain untested. The infection fatality rate (IFR) and infection hospitalization rate (IHR), calculated using the estimated total infections based on a representative sample of a population, is a better metric to assess the actual toll of the disease.

**Objective:** To determine the IHR and IFR for COVID-19 using the statewide SARS-CoV-2 seroprevalence estimates for the non-congregate population in Connecticut.

**Design:** Cross-sectional.

**Setting:** Adults residing in a non-congregate setting in Connecticut between March 1 and June 1, 2020.

**Participants:** Individuals aged 18 years or above.

**Exposure:** Estimated number of adults with SARS-CoV-2 antibodies.

**Main Outcome and Measures:** COVID-19-related hospitalizations and deaths among adults residing in a non-congregate setting in Connecticut between March 1 and June 1, 2020.

**Results:** Of the 2.8 million individuals residing in the non-congregate settings in Connecticut through June 2020, 113,515 (90% CI 56,758–170,273) individuals had SARS-CoV-2 antibodies. There were a total of 9425 COVID-19-related hospitalizations and 4071 COVID-19-related deaths in Connecticut between March 1 and June 1, 2020, of which 7792 hospitalizations and 1079 deaths occurred among the non-congregate population. The overall COVID-19 IHR and IFR was 6.86% (90% CI, 4.58%–13.72%) and 0.95% (90% CI, 0.63%–1.90%) among the non-congregate population. Older individuals, men, non-Hispanic Black individuals and those belonging to New Haven and Litchfield counties had a higher burden of hospitalization and deaths, compared with younger individuals, women, non-Hispanic White or Hispanic individuals, and those belonging to New London county, respectively.

**Conclusion and Relevance:** Using representative seroprevalence estimates, the overall COVID-19 IHR and IFR were estimated to be 6.86% and 0.95% among the non-congregate population in Connecticut. Accurate estimation of IHR and IFR among community residents is important to guide public health strategies during an infectious disease outbreak.

## BACKGROUND

Accurate estimation of the hospitalization and fatality rate is important to guide public health strategies during infectious disease outbreaks. Although the case fatality rate (CFR), defined as the proportion of deaths of the confirmed cases, is a commonly used metric, it will be biased based on the availability of testing, especially early in the outbreak.^1^ Moreover, since COVID-19 symptoms range widely, mild or asymptomatic infections may be untested. Thus, the number of infections confirmed by testing will underestimate the total infections, inflating the estimated fatality rate.

A better estimate of the actual toll is the infection fatality rate (IFR), defined as the proportion of deaths of the total infected individuals, in which the denominator is ideally based on a representative sample of a population. For hospitalizations, the infection hospitalization rate (IHR) is a comparable measure. Accordingly, using results of the recently conducted statewide SARS-CoV-2 seroprevalence study—the Post-Infection Prevalence study (PIP)—in Connecticut,^2^ we assessed the SARS-CoV-2 IHR and IFR.

## METHODS

Based on the PIP study,^2^ the seroprevalence of SARS-CoV-2 antibodies was 4.0% (90% CI 2.0%–6.0%) among a representative population of adults residing in non-congregate settings in Connecticut before June 2020. We used this estimate to calculate the overall population estimates for individuals infected with SARS-CoV-2 using the 2018 American Community Survey. We also estimated the number of individuals with SARS-CoV-2 antibodies by age, sex, race/ethnicity, and region.

Information on the total COVID-19-related hospitalizations and deaths among the non-congregate population in Connecticut, between March 1 and June 1, 2020, was provided by the Connecticut Hospital Association and the Connecticut Department of Public Health, respectively. The diagnostic codes used to identify COVID-19-related hospitalizations are listed in **eTable 1**. Total COVID-19 deaths included both confirmed and probable COVID-19 deaths (details in **eMethods**).

**Table 1.** Infection hospitalization rate and infection fatality rate among the non-congregate population in Connecticut between March 1 and June 1, 2020, by sociodemographic characteristics.

|  | **COVID-19-related Hospitalizations, N** | **Infection Hospitalization Rate, Weighted % (90% CI)** | **Total\* COVID-19-related Deaths, N** | **Total Infection Fatality Rate, Weighted % (90% CI)** |
| --- | --- | --- | --- | --- |
| **State Total** | 7792 | 6.86 (4.58–13.72) | 1079 | 0.95 (0.63–1.90) |
| **Sex** |  |  |  |  |
| Men | 4166 | 12.21 (6.23–NE) | 815 | 2.39 (1.22–59.71) |
| Women | 3625 | 4.64 (3.00–10.25) | 264 | 0.34 (0.22–0.75) |
| **Age-group** |  |  |  |  |
| 18-29 | 288 | 0.80 (0.36–NE) | 5 | 0.01 (0.01–NE) |
| 30-44 | 855 | 2.68 (1.38–43.85) | 34 | 0.11 (0.06–1.74) |
| 45-54 | 1013 | 3.09 (1.81–10.74) | 59 | 0.18 (0.11–0.63) |
| 55-64 | 1660 | 12.43 (6.46–NE) | 174 | 1.30 (0.68–16.94) |
| ≥65 | 3918 | 79.89 (31.96–NE) | 807 | 16.46 (6.58–NE) |
| **Race/Ethnicity** |  |  |  |  |
| Non-Hispanic Black | 1762 | 9.90 (5.32–70.40) | 259 | 1.46 (0.78–10.35) |
| Non-Hispanic White | 3948 | 7.42 (4.56–20.05) | 593 | 1.12 (0.68–3.01) |
| Hispanic | 845 | 1.04 (0.78–1.57) | 190 | 0.23 (0.17–0.35) |
| **County** |  |  |  |  |
| New Haven | 2101 | 9.04 (4.65–NE) | 270 | 1.16 (0.60–19.74) |
| New London | 148 | 4.04 (1.37–NE) | 21 | 0.57 (0.19–NE) |
| Middlesex | 231 | NE | 32 | NE |
| Fairfield | 2665 | 6.39 (3.28–NE) | 399 | 0.96 (0.49–18.16) |
| Hartford | 1755 | 6.21 (3.27–62.08) | 294 | 1.04 (0.55–10.40) |
| Litchfield | 195 | 8.26 (2.75–NE) | 39 | 1.65 (0.55–NE) |
| Abbreviations: CI, confidence interval; NE, non-estimable |  |  |  |  |
| *Total COVID-19-related deaths includes both confirmed and probable COVID-19 deaths. |  |  |  |  |

IHR and IFR were defined as the number of individuals who were hospitalized and died, respectively, due to COVID-19 divided by the total estimated number of individuals who had COVID-19 using the seroprevalence estimates. The margin of error for our estimates were calculated at the 90% confidence level in accordance with the design of the PIP study, however, estimates at 95% CI have also been provided. Due to sample size limitations, the upper end of the confidence interval was non-estimable (NE) when stratifying by some of the sociodemographic characteristics. All statistical analyses were performed using R version 4.0.2.

## RESULTS

Of the 2.8 million individuals residing in the non-congregate settings in Connecticut, 113,515 (90% CI 56,758–170,273) had SARS-CoV-2 antibodies (**eTable 2**). Between March 1 and June 1, 2020, there were a total of 9425 COVID-19-related hospitalizations and 4071 COVID-19-related deaths in Connecticut, of which 7792 hospitalizations and 1079 deaths occurred among the non-congregate population.

The overall COVID-19 IHR and IFR was 6.86% (90% CI, 4.58%–13.72%) and 0.95% (90% CI, 0.63%–1.90%) among the non-congregate population (**Table 1)**. When compared with women, men had a higher IHR (12.21% [90% CI, 6.23%–NE] vs 4.64% [90% CI, 3.00%– 10.25%]) and IFR (2.39% [90% CI, 1.22%–59.71%] vs 0.34% [90% CI, 0.22%–0.75%]). By age, IHR and IFR ranged from 0.80% (90% CI, 0.36%–NE) and 0.01% (90% CI, 0.01%–NE) among individuals aged 18–29 years, to 12.43% (90% CI, 6.46%–NE) and 1.30% (90% CI, 0.68%–16.94%) among those aged 55-64 years. By race/ethnicity, IHR and IFR were significantly higher among Black individuals (9.90% [90% CI, 5.32%–70.40%] and 1.46% [90% CI, 0.78%–10.35%]), as compared with White (7.42% [90% CI, 4.56%–20.05%] and 1.12% [90% CI, 0.68%–3.01%]) and Hispanic (1.04% [90% CI, 0.78%–1.57%] and 0.23% [90% CI, 0.17%–0.35%]) individuals. New Haven and Litchfield counties had the highest IHR (9.04% [90% CI, 4.65%–NE]) and IFR (1.65% [90% CI, 0.55%–NE]), respectively, whereas New London county had the lowest IHR (4.04% [90% CI, 1.37%–NE]) and IFR (0.57% [90% CI, 0.19%–NE]). IHR and IFR estimates with 95% CI are presented in **eTable 3**. The confirmed and probable IFRs are shown separately in **eTable 4**.

## DISCUSSION

Using seroprevalence estimates, we found that, through June 2020, Connecticut’s COVID-19 IHR and IFR were 6.86% and 0.95%, respectively. The rates of hospitalization and death varied and older individuals, men, non-Hispanic Black individuals, and those belonging to New Haven and Litchfield counties had the highest burden of adverse outcomes. Our estimates are distinctive because they reflect people living in the community and are based on a methodology that sought to obtain a representative estimate for the denominator.

There has been continued controversy about the IFR and the literature is replete with widely varied estimates.^3-7^ A recent meta-analysis on COVID-19 IFR, estimated a global pooled COVID-19 IFR of 0.68%,^5^ with values ranging from 0.01% to 1.60%. However, the studies included did not have a representative sample or separate out special populations, such as those in nursing homes. Moreover, IFR is not an inherent characteristic of the disease, but rather a confluence of the pathogen virulence, sociodemographic and clinical characteristics of the population, health care availability and quality, therapeutic availability, and accurate counting and reporting of COVID-19-related deaths. As such, an overall IFR may not be very informative given the heterogeneity among subgroups.

The COVID-19 IHR is not well described and most studies report the CHR, which also varies widely in the literature.^8-10^ The CDC estimated a US COVID-19 CHR of 14.0% for infections before June 2020.^10^ As expected, this CHR estimate was higher than Connecticut’s estimated IHR in our study (6.9%), as IHR includes the total estimated infections rather than detected positive cases only. Moreover, our estimate excluded individuals from congregate settings, which had a higher burden of adverse outcomes.

Our subgroup findings are notable. Although it has been noted that age and sex are associated with disease severity, prior studies have been hampered by including nursing home residents and biased by testing patterns. We had the opportunity to identify hospitalizations and deaths among the non-congregate population in Connecticut and show that, even in the community, these associations remain. Of note, Black individuals had a higher IFR and IHR than White individuals, though the difference was not statistically significant. Prior studies have shown that Black individuals have had disproportionately higher infection rates, even as some studies indicate that hospital mortality does not vary by race/ethnicity.^11^ Our findings highlight that the burden of COVID-19 among Black subpopulations is not just about infection rates but also worse outcomes. The IHR for Hispanic individuals was lower than for White and Black individuals, which was in accordance with previously reported low hospital admission rates among Hispanic individuals testing positive for SARS-CoV-2 in Baltimore-Washington, DC^12^ and may be associated with the lower insurance rates among the Hispanic subpopulation in Connecticut.^13^

Our study has some limitations. First, total COVID-19-related hospitalizations could have been underestimated due to limited testing availability, underestimating the IHR estimate. Second, although antibodies are specific indicators of past SARS-CoV-2 infection, their concentration may decrease few months after exposure^14^ and there are differences in the sensitivity of available assays.^15^ Hence, the total infections may be biased lower, overestimating our estimates. Nevertheless, representative seroprevalence studies provide important information regarding infections in a community and can provide robust estimates of the IHR and IFR, when combined with hospitalization and death data.

In conclusion, using representative seroprevalence estimates, we estimate an IHR and IFR of 6.86% (90% CI 4.58%–13.72%) and 0.95% (90% CI 0.63%–1.90%), respectively, for COVID-19 infections through June 1, 2020, among the non-congregate population in Connecticut.

## Data Availability

N/A

## ACKNOWLEDGEMENTS

The authors thank the Connecticut Department of Public Health and the Connecticut Hospital Association for their support.

