## Supplemental Material for "SARS-CoV-2 Infection Hospitalization Rate and Infection Fatality Rate among the Non-Congregant Population in Connecticut"

**Table of contents:**

eMethods. COVID-19 case classification criteria

eTable 1. ICD-10 codes to identify COVID-19 visits by Connecticut Hospital Association.

eTable 2. Estimated number of individuals with SARS-CoV-2-specifc antibodies among the non-congregate population in Connecticut, between March 1 and June 1, 2020, by sociodemographic characteristics.

eTable 3. Estimates for infection hospitalization rate and infection fatality rate at 95% margin of error among the non-congregate population in Connecticut between March 1 and June 1, 2020, by sociodemographic characteristics

eTable 4. Confirmed, probable, and total infection fatality rate among the non-congregate population in Connecticut between March 1 and June 1, 2020, by sociodemographic characteristics.

**eMethods. COVID-19 case classification criteria**

Case classifications –

- Confirmed:
  - Meets confirmatory laboratory evidence.
- Probable:
  - Meets clinical criteria AND epidemiologic evidence with no confirmatory laboratory testing performed for COVID-19.
  - Meets presumptive laboratory evidence AND either clinical criteria OR epidemiologic evidence.
  - Meets vital records criteria with no confirmatory laboratory testing performed for COVID19.

**Source:** Standardized surveillance case definition and national notification for 2019 novel coronavirus disease (COVID-19). Council of State and Territorial Epidemiologists. https://cdn.ymaws.com/www.cste.org/resource/resmgr/2020ps/interim-20-id-01_covid-19.pdf. Published 2020. Accessed September 8, 2020.

**eTable 1. ICD-10 codes to identify COVID-19 visits by Connecticut Hospital Association.**

| ICD-10 Codes | Description |
|---|---|
| J1289 and (B9729 or U071) | COVID-related Pneumonia |
| (J208 or J40) and (B9729 or U071) | COVID-related Bronchitis |
| (J988 or J22) and (B9729 or U071) | COVID-related respiratory infection |
| J80 and (B9729 or U071) | COVID-related ARDS |
| U071 | COVID-19 |

Note: the above diagnosis codes can be in any position, in other words, they do not have to be the principle diagnosis code.

**eTable 2. Estimated number of individuals with SARS-CoV-2-specifc antibodies among the non-congregate population in Connecticut, between March 1 and June 1, 2020, by sociodemographic characteristics.**

| Characteristics | Total Population*, N | Seroprevalence† of SARS-CoV-2 antibodies, % (±MOE at 90% CI) | Estimated number of individuals with antibodies, N (90% CI) | Seroprevalence† of SARS-CoV-2 antibodies, % (±MOE at 95% CI) | Estimated number of individuals with antibodies, N (95% CI) |
|---|---|---|---|---|---|
| **State total** | 2,837,877 | 4.0% (±2.0) | 113,515 (56,758 – 170,273) | 4.0% (±2.3) | 113,515 (48,244 – 178,786 |
| **Sex** | | | | | |
| Men | 1,365,019 | 2.5% (±2.4%) | 34,125 (1,365 – 66,886) | 2.5% (±2.9%) | 34,125 (NE – 73,711) |
| Women | 1,472,858 | 5.3% (±2.9%) | 78,061 (35,349 – 120,774) | 5.3% (±3.5%) | 78,061 (26,511– 129,612) |
| **Age group** | | | | | |
| 18-29 | 564,738 | 6.4% (±7.7%) | 36,143 (NE – 79,628) | 6.4% (±9.2%) | 36,143 (NE – 88,099) |
| 30-44 | 649,874 | 4.9% (±4.6%) | 31,844 (1,950 – 61,738) | 4.9% (±5.5%) | 31,844 (NE – 67,587) |
| 45-54 | 496,628 | 6.6% (±4.7%) | 32,777 (9,436 – 56,119) | 6.6% (±5.6%) | 32,777 (NE – 60,589) |
| 55-64 | 513,656 | 2.6% (±2.4%) | 13,355 (1,027 – 25,683) | 2.6% (±2.9%) | 13,355 (NE – 28,251) |
| ≥65 | 612,981 | 0.8% (±1.2%) | 4,904 (NE – 12,260) | 0.8% (±1.4%) | 4,904 (NE – 13,486) |
| **Race/Ethnicity** | | | | | |
| Non-Hispanic Black | 278,112 | 6.4% (±5.5%) | 17,799 (2,503 – 33,095) | 6.4% (±6.5%) | 17,799 (NE – 35,876) |
| Non-Hispanic White | 1,969,487 | 2.7% (±1.7%) | 53,176 (19,695 – 86,657) | 2.7% (±2.0%) | 53,176 (13,786 – 92,566) |
| Hispanic | 408,654 | 19.9% (±6.7%) | 81,322 (53,942 – 108,702) | 19.9% (±8.0%) | 81,322 (48,630 – 114,015) |
| **County** | | | | | |
| New Haven | 683,928 | 3.4% (±3.2%) | 23,254 (1,368 – 45,139) | 3.4% (±3.8%) | 23,254 (NE – 49,243) |
| New London | 215,679 | 1.7% (±3.3%) | 3,667 (NE – 10,784) | 1.7% (±4.0%) | 3,667 (NE – 12,294) |
| Middlesex | 133,380 | - | - | - | - |
| Fairfield | 732,172 | 5.7% (±5.4%) | 41,734 (2,197 – 81,271) | 5.7% (±6.4%) | 41,734 (NE – 88,593) |
| Hartford | 706,631 | 4.0% (±3.6) | 28,265 (2,827 – 53,704) | 4.0% (±4.3%) | 28,265 (NE – 58,650) |
| Litchfield | 147,570 | 1.6% (±3.2%) | 2,361 (NE – 7,083) | 1.6% (±3.8%) | 2,361 (NE – 7,969) |

* Source: 2018 American Community Survey

† Source: Post-Infection Prevalence Study

Abbreviations: CI, confidence interval; MOE, margin of error

**eTable 3. Estimates for infection hospitalization rate and infection fatality rate at 95% margin of error among the non-congregate population in Connecticut between March 1 and June 1, 2020, by sociodemographic characteristics**

| | COVID-19-related Hospitalizations, N | Infection Hospitalization Rate, Weighted % (95% CI) | Total* COVID-19-related Deaths, N | Total Infection Fatality Rate, Weighted % (95% CI) |
|---|---|---|---|---|
| **State Total** | 7792 | 6.86 (4.36–16.15) | 1079 | 0.95 (0.60–2.24) |
| **Sex** | | | | |
| Men | 4166 | 12.21 (5.67–NE) | 815 | 2.39 (1.11–NE) |
| Women | 3625 | 4.64 (2.80–14.21) | 264 | 0.34 (0.20–1.00) |
| **Age-group** | | | | |
| 18-29 | 288 | 0.80 (0.33–NE) | 5 | 0.01 (0.01–NE) |
| 30-44 | 855 | 2.68 (1.27–NE) | 34 | 0.11 (0.05–NE) |
| 45-54 | 1013 | 3.09 (1.67–NE) | 59 | 0.18 (0.10–NE) |
| 55-64 | 1660 | 12.43 (5.88–NE) | 174 | 1.30 (0.62–NE) |
| ≥65 | 3918 | 79.89 (29.05–NE) | 807 | 16.46 (5.98–NE) |
| **Race/Ethnicity** | | | | |
| Non-Hispanic Black | 1762 | 9.90 (1.90–NE) | 259 | 1.46 (0.72–NE) |
| Non-Hispanic White | 3948 | 7.42 (11.00–28.64) | 593 | 1.12 (0.64–4.30) |
| Hispanic | 845 | 1.04 (0.74–1.74) | 190 | 0.23 (0.17–0.39) |
| **County** | | | | |
| New Haven | 2101 | 9.04 (4.27–NE) | 270 | 1.16 (0.55–NE) |
| New London | 148 | 4.04 (1.20–NE) | 21 | 0.57 (0.17–NE) |
| Middlesex | 231 | NE | 32 | NE |
| Fairfield | 2665 | 6.39 (3.01–NE) | 399 | 0.96 (0.45–NE) |
| Hartford | 1755 | 6.21 (2.99–NE) | 294 | 1.04 (0.50–NE) |
| Litchfield | 195 | 8.26 (2.45–NE) | 39 | 1.65 (0.49–NE) |

Abbreviations: CI, confidence interval; NE, non-estimable

*Total COVID-19-related deaths includes both confirmed and probable COVID-19 deaths.

**eTable 4. Confirmed, probable, and total infection fatality rate among the non-congregate population in Connecticut between March 1 and June 1, 2020, by sociodemographic characteristics.**

| | Confirmed deaths, N | Confirmed IFR, weighted % (90% CI) | Confirmed IFR, weighted % (95% CI) | Probable deaths, N | Probable IFR, weighted % (90% CI) | Probable IFR, weighted % (95% CI) |
|---|---|---|---|---|---|---|
| **State Total** | 860 | 0.76 (0.51–1.52) | 0.76 (0.48–1.78) | 219 | 0.19 (0.13–0.39) | 0.19 (0.12–0.45) |
| **Sex** | | | | | | |
| Men | 598 | 1.75 (0.89–43.81) | 1.75 (0.81–NE) | 217 | 0.64 (0.32–15.90) | 0.64 (0.29–NE) |
| Women | 262 | 0.34 (0.22–0.74) | 0.34 (0.20–0.99) | 2 | 0.00 (0.00–0.01) | 0.00 (0.00–0.01) |
| **Age group** | | | | | | |
| 18-29 | 5 | 0.01 (0.01–NE) | 0.01 (0.01–NE) | 0 | 0.00 (0.00–0.00) | 0.00 (0.00–0.00) |
| 30-44 | 30 | 0.09 (0.05–1.54) | 0.09 (0.04–NE) | 4 | 0.01 (0.01–0.21) | 0.01 (0.01–NE) |
| 45-54 | 47 | 0.14 (0.08–0.50) | 0.14 (0.08–NE) | 12 | 0.04 (0.02–0.13) | 0.04 (0.02–NE) |
| 55-64 | 148 | 1.11 (0.58–14.41) | 1.11 (0.52–NE) | 26 | 0.19 (0.10–2.53) | 0.19 (0.09–NE) |
| ≥65 | 630 | 12.85 (5.14–NE) | 12.85 (4.67–NE) | 177 | 3.61 (1.44–NE) | 3.61 (1.31–NE) |
| **Race/ethnicity** | | | | | | |
| Non-Hispanic Black | 220 | 1.24 (0.66–8.79) | 1.24 (0.61–NE) | 39 | 0.22 (0.12–1.56) | 0.22 (0.11–NE) |
| Non-Hispanic White | 450 | 0.85 (0.52–2.28) | 0.85 (0.49–3.26) | 143 | 0.27 (0.17–0.73) | 0.27 (0.15–1.04) |
| Hispanic | 160 | 0.20 (0.15–0.30) | 0.20 (0.14–0.33) | 30 | 0.04 (0.03–0.06) | 0.04 (0.03–0.06) |
| **County** | | | | | | |
| New Haven | 236 | 1.01 (0.52–17.25) | 1.01 (0.48–NE) | 34 | 0.15 (0.08–2.49) | 0.15 (0.07–NE) |
| New London | 15 | 0.41 (0.14–NE) | 0.41 (0.12–NE) | 6 | 0.16 (0.06–NE) | 0.16 (0.05–NE) |
| Middlesex | 26 | NE | NE | 6 | NE | NE |
| Fairfield | 315 | 0.75 (0.39–14.34) | 0.75 (0.36–NE) | 84 | 0.20 (0.10–3.82) | 0.20 (0.09–NE) |
| Hartford | 224 | 0.79 (0.42–7.92) | 0.79 (0.38–NE) | 70 | 0.25 (0.13–2.48) | 0.25 (0.12–NE) |
| Litchfield | 28 | 1.19 (0.40–NE) | 1.19 (0.35–NE) | 11 | 0.47 (0.16–NE) | 0.47 (0.14–NE) |

Abbreviations: CI, confidence interval; IFR, infection fatality rate; NE, non-estimable
